# Co-detection of respiratory pathogens in patients hospitalized with Coronavirus viral disease-2019 pneumonia

**DOI:** 10.1101/2020.04.06.20052522

**Authors:** María Luisa Blasco, Javier Buesa, Javier Colomina, María José Forner, María José Galindo, Jorge Navarro, José Noceda, Josep Redón, Jaime Signes-Costa, David Navarro

## Abstract

There is scarce information on the frequency of co-detection of respiratory pathogens (RP) in patients with Covid-19. Documentation of coinfections in Covid-19 pneumonia patients may be relevant for appropriate clinical and therapeutic management of patients. Between March 4th and March 28th, 2020, a total of 183 adult patients testing positive by SARS CoV-2 RT-PCR on respiratory specimens were hospitalized with interstitial pneumonia at our center, of whom 103 were tested for other RP by a multiplexed PCR assay. Three patients had a positive result for either one (n=2; Coronavirus HKU1 or Mycoplasma pneumoniae) or two targets (n=1; Influenza virus A (H3) and Respiratory syncytial virus B). Twenty-three patients testing negative by SARS CoV-2 RT-PCR and presentig with clinical, laboratory findings and imaging compatibe with Covid-19 pneumonia underwent RP screening. Of these, 6 (26%) had a positive result for a single RP. Our data indicate that despite the apparent rarity of coinfections in patients with Covid-19 pneumonia, routine testing for RP should be advised, since agents for which specific therapy can be prescribed may be detected.

---

Clinical, laboratory and imaging characteristics of Coronavirus disease 2019 (Covid-19) and risk factors associated with poor outcomes have been reported in various studies (1-5). Information on whether other respiratory pathogens (RP) were co-detected in these patients was not provided, despite the fact that coinfection with RP has been reported to occur in this clinical setting (6-9). Documentation of coinfections in Covid-19 pneumonia patients may be relevant not only for appropriate clinical and therapeutic management of patients, but also to precisely characterize disease features and delineate risk factors potentially impacting on clinical outcomes. Here, we report on our experience on this topic, gathered between March 4th and March 28th, 2020, at the Clinic University Hospital of Valencia, a tertiary teaching hospital with 586 beds which serves Clínico-Malvarrosa Health Department (attending 368.000 inhabitants in the northeast of the city). The study was approved by the Ethical Committee of University Clinic Hospital, INCLIVA, Valencia. Informed consent was not requested as laboratory analyses reported herein were conducted routinely in our patients following local guidelines.

A total of 183 adult patients testing positive by SARS CoV-2 RT-PCR on respiratory specimens were hospitalized with interstitial pneumonia, of whom 103 (64 males/39 females; median age, 64 years; range, 19-100 years) were tested for other RP by a multiplexed PCR assay (10) (Table 1). Three patients (2.9%) had positive results for either one (n=2; Coronavirus HKU1 or *Mycoplasma pneumoniae)* or two targets (n=1; Influenza virus A (H3) and Respiratory syncytial virus B). Twenty-three patients (14 males/9 females; median age, 54 years; range, 31-79 years) testing negative by SARS CoV-2 RT-PCR in a single respiratory specimen, presentig with clinical, laboratory findings and imaging compatibe with Covid-19 pneumonia (1-5) underwent RP screening. Of these, 6 (26%) had a positive result for a single RP.

**TABLE 1.**
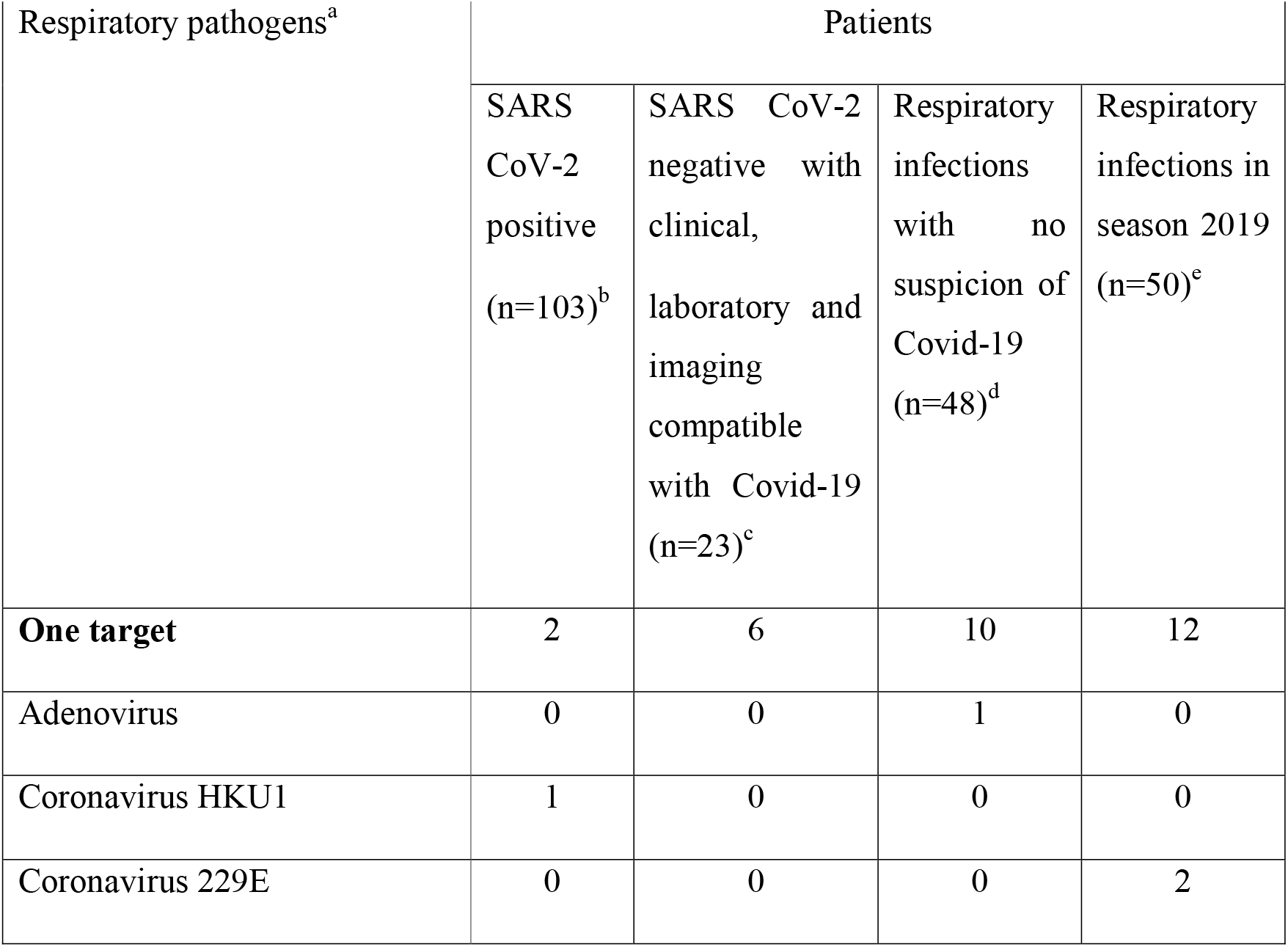
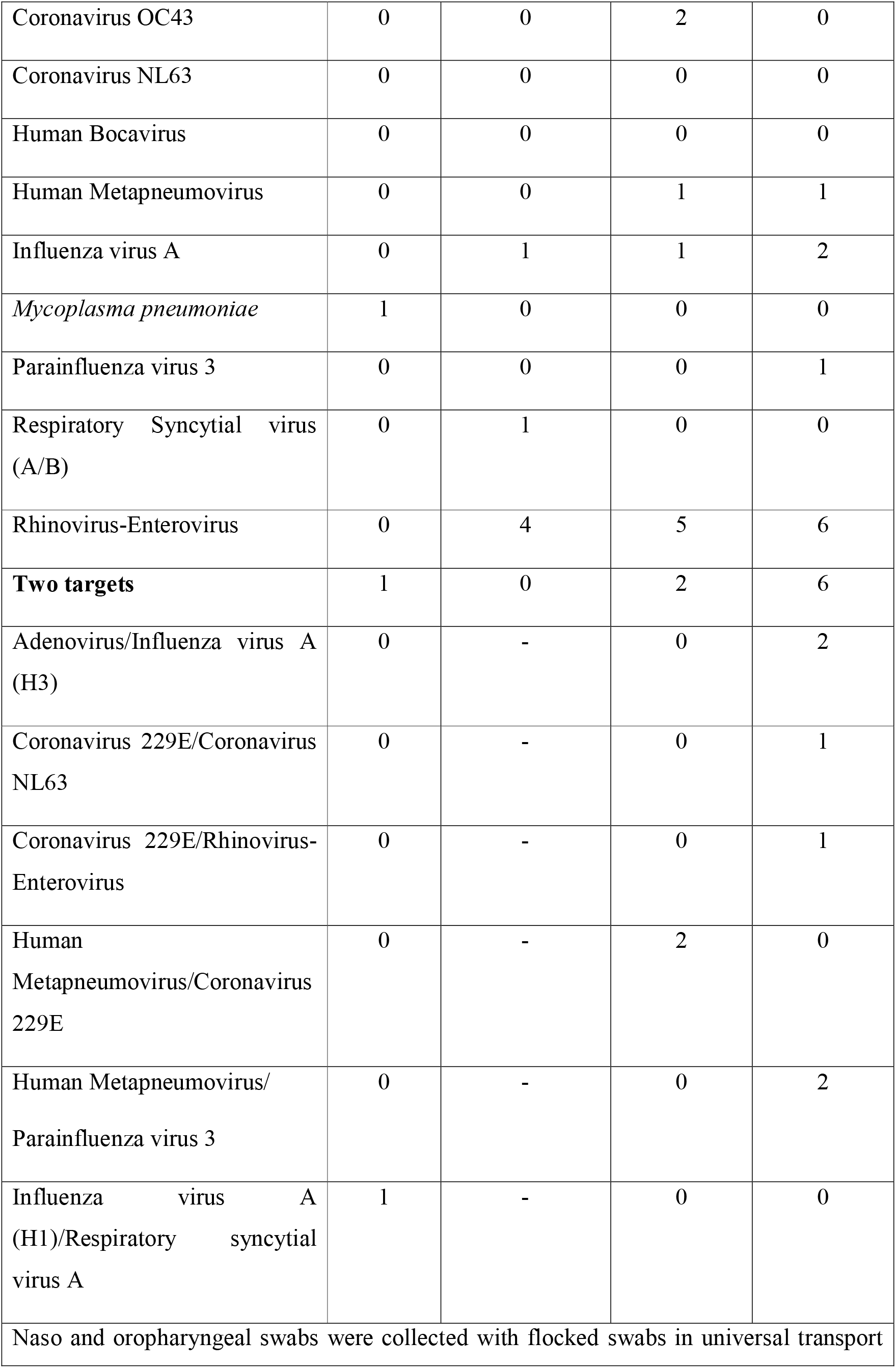
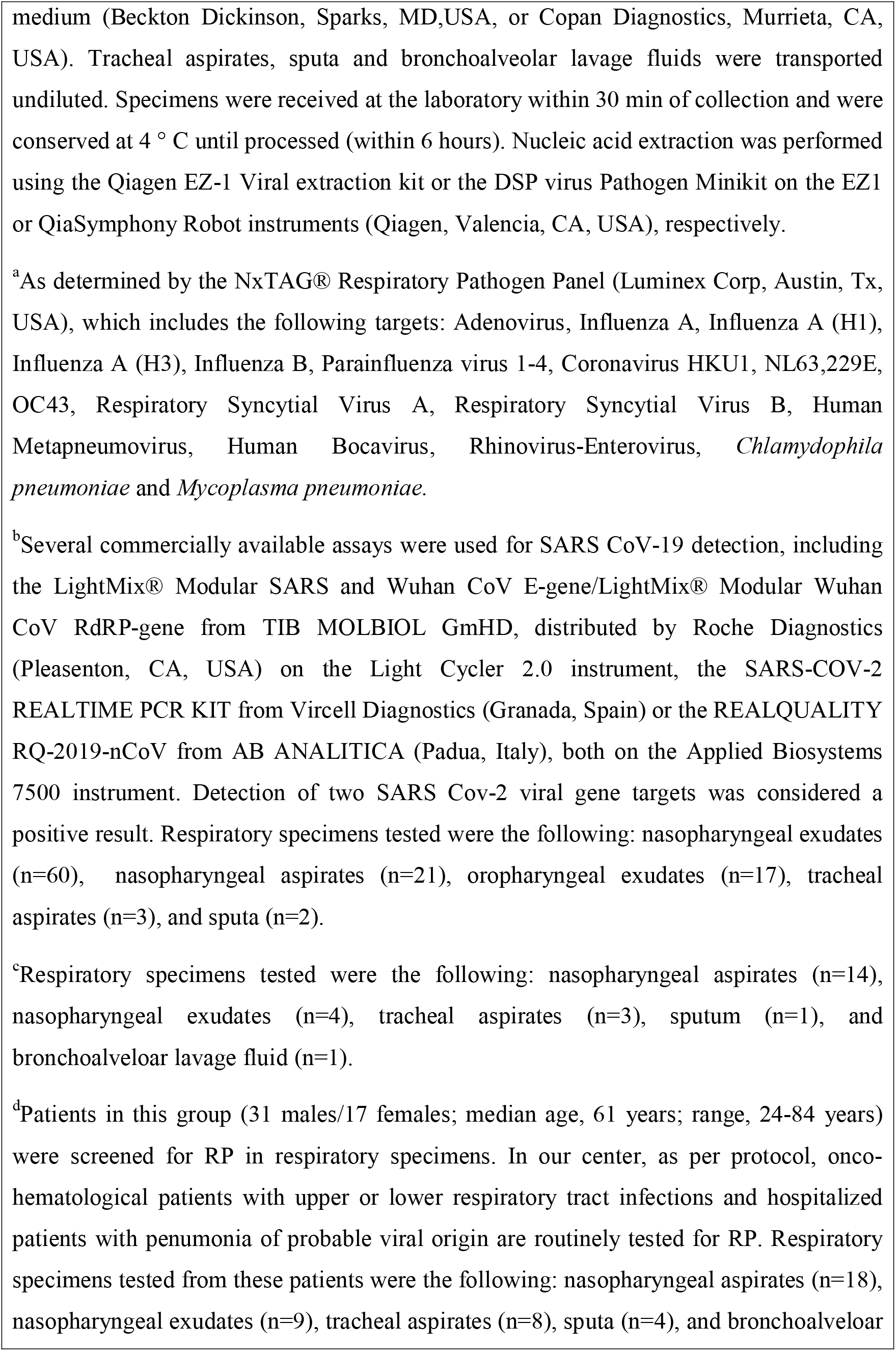
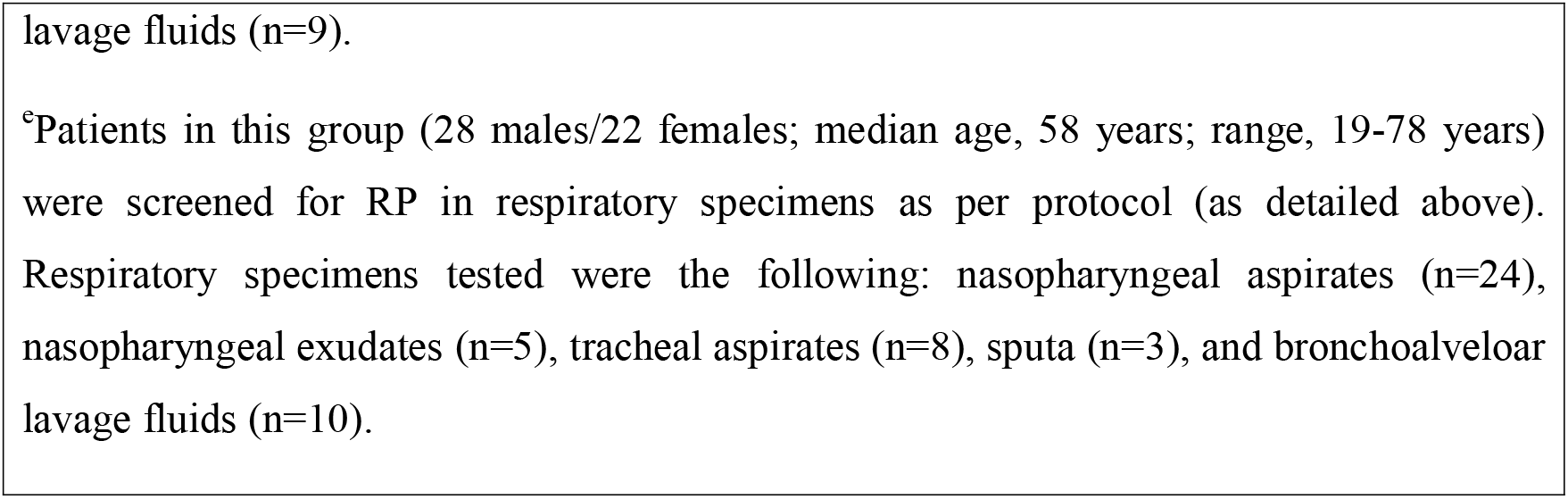
Detection of respiratory pathogens by multiplexed PCR in the study patients.

Coinfection with RP in adult patients with respiratory tract infections occurs commonly (11,12). In line with this, coinfection with RP was observed in 2 out of 12 patients (25%) presenting with upper or lower respiratory tract infections and no clinical suspicion of Covid-19 and testing positive for RP within the study period. The rate of co-detection of RP in the same time period of the preceding year was rather comparable (6 out of 18 patients testing positive for RP; 35%). Nevertheless, patients with Covid-19 pneumonia were found to be infrequently coinfected with other RP; this phenomenom could have been underestimated, since 26% of patients fulfilling criteria of this clinical entity (1-5) and testing negative by SARS CoV-2 RT-PCR, had RP detected in respiratory specimens. In this sense, false negative SARS CoV-2 PCR results in upper respiratory tract specimens occurs (13). The possibility of a viral interference phenomenon similar to that described for influenza virus A (sub)types should also be considered (14). Despite the apparent rarity of coinfections in patients with Covid-19 pneumonia, routine testing for RP should be advised, since agents for which specific therapy can be prescribed (i.e *Mycoplasma pneumoniae*, Influenza virus A, or Respiratory syncytial virus) may be detected. This approach may have a benefitial impact on patient survival.

## Data Availability

Due to the nature of this research, participants of this study did not agree for their data to be shared publicly, so supporting data is not available.

## Acknowledgements

We are grateful to all personnel who work at Clinic University Hospital for their dedicated and tireless effort in the fight against Covid-19.

